# Limited predictive value of the gut microbiome and metabolome for response to biological therapy in inflammatory bowel disease

**DOI:** 10.1101/2024.05.10.24307195

**Authors:** Femke M. Prins, Iwan J. Hidding, Marjolein A.Y. Klaassen, Valerie Collij, Johannes P.D. Schultheiss, Werna T.C. Uniken Venema, Amber Bangma, Jurne B. Aardema, Bernadien H. Jansen, Wout G.N. Mares, Ben J.M. Witteman, Eleonora A.M. Festen, Gerard Dijkstra, Marijn C. Visschedijk, Herma H. Fidder, Arnau Vich Vila, Bas Oldenburg, Ranko Gacesa, Rinse K. Weersma

**Author notes:** Corresponding author Rinse K. Weersma, MD, PhD, Department of Gastroenterology and Hepatology & Department of Genetics, University of Groningen and University Medical Center Groningen, PO Box 30.001, 9700RB Groningen, the Netherlands., 9306. Shared first author (Femke M. Prins, Iwan J. Hidding). Shared second author (Marjolein A.Y. Klaassen, Valerie Collij).

## Abstract

**Introduction:** The complexity of Inflammatory Bowel Diseases (IBD) presents challenges for the management of these diseases. Predicting treatment outcomes remains difficult, leading to suboptimal outcomes and high costs. Emerging evidence suggests the potential of the gut microbiome in predicting response to biologic treatments. In this prospective study we aimed to predict treatment response to vedolizumab and ustekinumab in 79 IBD patients by integrating clinical data, gut microbiome profiles and fecal metabolites and validating these findings in a replication cohort of 47 IBD patients.

**Methods:** Treatment response was defined as either continuation or discontinuation of the biologic at six months. We performed whole genome metagenomic shotgun sequencing on the baseline fecal samples to detect microbial and functional profiles. Additionally, over 1000 metabolites were captured through untargeted metabolomic profiling. Baseline diversity, dissimilarity and differential abundance analyses compared responder and non-responder groups. The prediction tool CoDaCoRe was used to identify predictive log-ratio biomarkers. We tested our identified ratios in an external cohort and attempted to replicate the microbiome-based signals of previous studies. Finally, we used a neural-network framework to model the relationship between metabolites and microbes, comparing these clusters in responders and non-responders and tested our approach with different definitions of response.

**Results:** We identified seven metabolites to be differentially abundant between responders and non-responders (FDR < 0.05). However, no significant differences in bacterial species and pathways were detected at baseline between responders and non-responders. Our prediction analysis indicated only marginal predictive utility of the gut microbiome and fecal metabolites for treatment response, when compared to a clinical model using fecal calprotectin, disease duration and disease activity, among other factors (AUC only clinical features: 0.71±0.13, AUC microbial and clinical features: 0.73±0.12). The main predictive features of these models were the disease activity and previous anti-TNF use combined with high abundance of *Phocaeicola vulgatus, Bacteroides uniformis* and *Alistipes onderdonkii,* and the low abundance of *Ruminococcus gnavus* and *Faecalibacterium prausnitzii.* Testing our identified ratio of 10 species in an external cohort of 47 IBD patients reinforced the lack of predictive power of the microbiome. No replication of previously published predictive signals of the microbiome was observed. Additionally, we identified 2 metabolite clusters and 1 microbiome cluster associated with response, and observed that predictors were highly dependent on the definition of response.

**Conclusion:** While previous studies of similar size have shown that microbial features can predict response to either vedolizumab (VedoNet), or vedolizumab, ustekinumab and anti-TNF, our comprehensive study found no significant differences in the gut microbiome at baseline between responders and non-responders among IBD patients treated with ustekinumab or vedolizumab. Microbial features added no predictive power to drug response in our IBD patient cohort or an independent replication cohort. Addition of metabolite features did not improve predictive power. These findings suggest minimal impact of the pre-treatment gut microbiome on treatment outcomes with these medications in IBD patients with long term chronic disease. Generalizability beyond initial study cohorts is limited, leaving predictors for individualized IBD-medication selection unidentified, but based on this work, likely do not lie in the fecal microbiome.

## Introduction

Inflammatory bowel diseases (IBD), including Crohn’s disease (CD) and ulcerative colitis (UC), are complex chronic inflammatory disorders of the gastrointestinal tract. They affect over 1.3 million individuals in Europe alone, posing significant challenges for gastroenterologists due to the considerable heterogeneity in onset and progression^1^. The precise etiology of IBD remains elusive, but it is currently believed that it involves a combination of genetic susceptibility, an inappropriate immune response to the gut microbiome, and a variety of environmental triggers^2^. This multifactorial nature adds complexity to managing IBD and hampers the development of potential therapeutic targets.

Traditionally, IBD treatment focused on reducing inflammation using immunomodulators with pleiotropic effects such as mesalazines, corticosteroids and azathioprine. Over the last two decades drugs that target specific components of the immune system became standard of care for induction and maintenance of remission in IBD, such as TNF-alpha inhibitors (e.g. infliximab), integrin receptor antagonists like vedolizumab (α4β7-integrin inhibitor) and inhibitors of IL-12/IL-23 (e.g. ustekinumab). Unfortunately, these drugs have hit a therapeutic ceiling and only reduce remission in one-half to two-third of patients^3,4^. Moreover, the ongoing necessity of surgical interventions^5^, the development of rare serious side effects, and high costs of these drugs, underscore the importance of identifying patient-specific characteristics to predict outcomes before treatment initiation.

Currently, there are no well-established biomarkers that predict whether a patient will respond to advanced therapy or not. Patient-related factors (such as age, sex, smoking habits) and disease-related factors (disease duration, location, activity), have not been found to be reliable in this respect^6^. Interestingly, longitudinal studies of the fecal microbiome have revealed differences in gut microbiome composition between patients with IBD who respond to biological treatments and those who do not. For instance, responders to TNF-alpha inhibitors tend to have higher abundances of certain bacteria like *Bifidobacterium*, *Clostridiales* and *Eubacterium rectale*^7,8^, while non-responders show a depletion of *Faecalibacterium prausnitzii*^9^. Microbial differences have also been observed for response to vedolizumab and ustekinumab^10,11^. Additionally, emerging evidence underscores the importance of metabolites derived from the gut microbiome in mediating host-microbiome interactions and immune responses^12^. Analyzing these metabolites offers valuable insights into the biochemical activity of the gut microbiome. A recent study demonstrated the predictive potential of these metabolites, showing that a specific ratio of metabolites can classify IBD and non-IBD samples^13^. Furthermore, recent studies showed that changes in bile acid levels are predictive of therapeutic response^14^, e.g. CD patients responding to TNF-alpha inhibitors exhibited higher serum levels of secondary bile acids^15^.

Considering the pivotal role of the gut microbiome and host immunity in the pathogenesis of IBD, we hypothesized that integrating clinical data, metagenomic gut microbiome profiles, and fecal metabolomics can predict response to biologics. To test this, we analyzed these data layers of 79 patients treated with either vedolizumab or ustekinumab and constructed response-prediction models.

## Methods

### Study cohort

A total of 100 patients were recruited for this study, with 50 patients initiating vedolizumab and 50 patients starting treatment with ustekinumab at the specialized IBD center of the Department of Gastroenterology and Hepatology of the University Medical Center Groningen (UMCG), the Netherlands. Inclusion criteria required patients to be 18 years or older and have a confirmed diagnosis of IBD for at least three months prior to inclusion, based on clinical, endoscopic, and histopathological criteria. Informed consent from all participants was obtained through Parelsnoer IRB (NL24572.018.08) and GEID (NL58808.042.16).

Clinical characteristics, fecal samples and lab results were collected at baseline, i.e. prior to the start of therapy. IBD-related clinical characteristics were derived from medical records, including the Montreal classification, IBD disease duration, prior medication use, and previous surgical interventions. Additionally, demographic and clinical features, such as age, sex, BMI, current medication use, smoking behavior, and disease severity scores (Harvey Bradshaw Index (HBI) for CD, Simple Clinical Colitis Activity Index (SCCAI) for UC), were determined at baseline. The decision to initiate vedolizumab or ustekinumab therapy was made by the patients’ treating physician.

### Defining response

Response to vedolizumab or ustekinumab was determined by drug survival (decision to continue the biologic treatment) at the six months mark, based on the physicians’ global assessments (including fecal calprotectin, serum CRP levels, and clinical disease activity scores). Consequently, if the treating physician discontinued the biologic treatment before six months, patients were classified as being non-responders. In addition, we performed analyses using other definitions of response since the definition of response lacks uniformity in the existing literature: i) sustained response, defined as the ongoing use of the biologic drug after two years, and ii) a reduction of ⩾3 points from baseline in HBI/SCCAI scores at 14 weeks for vedolizumab and 16 weeks for ustekinumab corresponding to a physician visit.

### Sample collection and processing

Patients produced and immediately froze the fecal samples at home using a provided stool collection kit. Research staff from the UMCG collected the feces on dry ice, using insulating polystyrene foam containers, and stored them at −80⁰C. DNA was extracted from fecal material using the QIAamp Fast DNA Stool Mini Kit and the QIAcube automated sample preparation system (Qiagen, Germany). The samples were sent to the sequencing facilities: vedolizumab samples were processed at NovoGene in Hong Kong, and ustekinumab samples were processed at Novogene in the United Kingdom (the different locations were due to the relocation of Novogene Europe). Both batches underwent whole genome shotgun metagenomic sequencing using the Illumina NovaSeq 6000 S4 flowcell with PE150. While no obvious batch effects were observed, we accounted for batch (biologic cohort) in our analyses. For the metabolomics, fresh frozen samples were drilled on dry ice until obtaining on average 0.5 mg of fecal material, transferred into a 2ml tube and shipped to Metabolon facilities for untargeted metabolomic measurements.

### Metagenomic and metabolomic data processing

First, we removed the Illumina adapters from the raw metagenomic reads and trimmed them using the *KneadData* (v0.12.0.) tool. We then removed reads aligning to the human genome using *Bowtie2* (v2.5.1). The taxonomic composition was profiled using *MetaPhlan4* (v4.0.6, library vOct22). This resulted in 2 kingdoms, 14 phyla, 151 classes, 170 orders, 206 families and 1087 species-level genome bins (SGBs), further referred to as species, for the total cohort. Bacterial species with a prevalence of more than 15% and a minimum mean relative abundance of 0.1% were kept for analyses (species n = 65) and taxa that were unclassified at species level were excluded. We profiled the abundance of microbial metabolic pathways using HUMAnN (v3.6), resulting in 476 predicted pathways. After applying filtering based on a prevalence of more than 15% and a minimum relative abundance of 0.001%, 193 pathways were left for analyses. Because of the compositional nature of the data, prior to statistical tests we transformed the microbial abundances and pathway abundances using the centered log-ratio method (CLR).

For the metabolomic data, raw data processing and quality control were performed according to Metabolon’s standards. This data was batch-normalized (raw values divided by the median of the samples in the batch) and missing values were imputed with the minimum value across all batches in the median scaled data. The data was then transformed using the natural log, since metabolomic data typically displays a log-normal distribution. For analyses we filtered the metabolites on presence in >70% of the samples (metabolites n = 816).

### Statistical analyses

Analyses were performed in R (v4.2.3) and Python (v3.8.8). All taxonomic microbiome analyses were performed at species level. The analyses were conducted for the total cohort and also stratified by biologic therapy.

#### Diversity and dissimilarity analyses

Alpha diversity was calculated using the Shannon index, a metric encompassing both species evenness and richness, using the *diversity* function of the R *vegan* package (v2.6-4). Differences between responders and non-responders were tested with the Wilcoxon rank-sum test. For beta-diversity, measuring the dissimilarity of microbial composition between samples, Aitchison distances (Euclidean distance of CLR transformed data) were computed using the function *vegdist* and visualized using Principal Coordinate Analysis (PCoA) plots. To test differences in community structure between responders and non-responders and the effect on variation in microbial and pathway composition, we performed a PERMANOVA test with 1000 permutations, as implemented in the *adonis2* function where we added sex, age, BMI and sequencing read depth and additional covariates known to affect the composition of the microbiome: proton-pump inhibitor (PPI) use, antibiotics use three months prior to sampling and history of bowel resections. The *betadisper* function was used to confirm whether outcomes of the PERMANOVA were influenced by variations in dispersion between groups.

#### Differential abundance of species, pathways and metabolites

We aimed to identify microbial species, pathways and metabolites whose baseline abundances differed between responders and non-responders. Differential abundance analysis was conducted using linear regression, considering age, sex, BMI, PPI-use, antibiotics use, history of bowel resections [yes/no], IBD diagnosis subtype, and sequencing read depth as covariates. Analyses were corrected for multiple testing by applying the Benjamini-Hochberg procedure, with significance defined as a false discovery rate (FDR) of <0.1. For the metabolites, using imputed peak area data, we analyzed bile acids and tested the logratio of primary bile acids (PBA) and secondary bile acids between responders and non-responders.

#### Metabolite-microbiome interaction network

To identify any metabolite-microbiome interactions and if any of these interactions are associated with responders or non-responders, we used MiMeNet (v1.0.0). As input we provided the metabolite raw AUC values and the microbiome relative abundances multiplied by the read counts of each file for pseudocounts. Additionally, we put the labels of responder or non-responder on each patient. We used the R iGraph package (v2.0.3) for visualizing the interaction graph.

#### Prediction of response outcomes

We used the CoDaCoRe package (v0.0.4) as our primary method to predict response to biologic therapy. This algorithm is designed to identify predictive log-ratio biomarkers in high-throughput sequencing data. The predictions were carried out independently for metagenomic data (taxonomy and pathways), metabolite data, clinical data and combinations of these data layers. For the taxonomic data, the input for the prediction models was the non-transformed species data with the same filtering as for the microbiome analyses (prevalence >15% samples and a minimum mean relative abundance of 0.1). The relative abundance was then multiplied by the aligned reads to generate pseudocounts per bacterial species. For the pathway prediction input we used similar preprocessing steps as for the bacterial species, first we filter on prevalence >0.15 and a minimum mean relative abundance of 0.001, then we multiply the leftover pathways by the reads in the processed samples to create pseudocounts as input for the CoDaCoRe models. For the metabolite input, raw AUC data was used. We predicted responses using the *logratiotype* setting *amalgamations,* i.e ratio constructed from the sum of features, and a lambda of 2. All predictions involved splitting samples into 75% training and 25% testing sets. Because of limited sample size, this step was permuted 100 times to obtain accurate values for the model AUC and test AUC. We summarized the features selected in the highest AUC ratio from CoDaCoRe for each permutation as a fraction of the total number of permutations. To determine the main predictors in each model, we applied a cutoff of 10% presence across all predictions. We then created ratios using the features chosen for the numerator and denominator and applied these ratios to the samples to visualize the difference between groups.

#### Prediction model performances

To assess the performance of the ratios, first, we created a Generalized Linear Model (GLM) based only on clinical factors. As input for the clinical data we used sex, BMI, age, fecal calprotectin, CRP, previous use of anti-TNF medication, disease duration, and disease activity. Disease activity was determined based on the baseline HBI and SCCAI scores (no activity HBI <5 or SCCAI <3, mild disease activity HBI 5-7 or SCCAI 3-5, moderate disease activity HBI 8-16 or SCCAI 6-11, and severe disease activity HBI >16 or SCCAI >11)^16^. We trained the model on 75% of the data, and then tested it on the remaining 25% of the data with 100 permutations to determine model AUC. Then we added the created ratio based on microbial features and compared performance. This was repeated with the ratio based on metabolite features, and then also with the ratio based on pathway features. We then combined all these ratios in one model to compare the full predictive power of our features compared to the model containing only clinical features.

### Replication efforts

#### Replication cohort

To validate the predictive features we identified in our cohort, we used a replication cohort combined from patients of a tertiary referral center (University Medical Center Utrecht) and a general hospital (Gelderse Vallei hospital). All patients had Crohn’s disease and started biologic therapy (vedolizumab, infliximab or adalimumab). Consent from all participants was obtained (METC 16-137). Inclusion period was between 2016 and 2020 The decision to initiate vedolizumab, infliximab or adalimumab therapy was made by the patients’ treating physician. Clinical characteristics and fecal samples were collected at baseline. Fecal samples were stored within 24 hours at −80⁰C, IBD related clinical characteristics were derived from medical records, including the Montreal classification, IBD disease duration, prior medication use, and previous surgical interventions. Additionally, demographic and clinical features, such as age, sex, BMI, current medication use, smoking behavior, and HBI were recorded at baseline.

As this cohort contained some overlapping biologic use (vedolizumab) and some non-overlapping biologic use (infliximab, adalimumab), we tested two different setups. First, we visualized the predictive features from our vedolizumab only subset in the vedolizumab patients of the Utrecht cohort. Second, we visualized the features from our total cohort in the anti-TNF subgroup of the Utrecht cohort. Subsequently we created GLMs testing the model performance based on the same clinical features used in our own cohort prediction, and then the same model including the microbiome ratio. We trained the vedolizumab only model on all our vedolizumab patients and tested this on the Utrecht vedolizumab patients. The anti-TNF model was trained on our full cohort and tested on the Utrecht anti-TNF patients.

#### VedoNet prediction

To test the previously described VedoNet prediction model^10^, we identified the metagenomic and clinical features (n=49) in our data overlapping with their features as input for a Sklearn (v.1.3.1) Random Forest model (1000 trees, depth of 45), using 4 split 5 repeats k-crossfold validation. Using only the samples from patients starting with vedolizumab, and after removing any samples with data missing for any of these features, we used 22 patients (13 responders and 9 non-responders) for this comparison.

#### Enterotypes

To replicate the prediction based on enterotypes^17^, we used Dirichlet Multinomial Mixtures (DMM) from the *DirechletMultinomial* package to determine enterotypes (community types). Community typing was performed on a genus-abundance matrix including all available study samples. We also performed DMM analysis on a combined matrix of samples from this study and 8298 samples from the Dutch Microbiome Project to improve accuracy of community typing. We looked at potential associations between response and patients baseline characteristics (sex, age, BMI, CRP, fecal calprotectin, smoking status, disease duration, previous resections, prior use of anti-TNF therapy, and enterotype). These variables were modeled as single explanatory variables in a logistic regression (*glm* function, family = binomial(link = “logit”)).

## Results

## 1. Cohort overview

### 1.1 Patient inclusion

A total of 100 patients were recruited (50 for each biologic). Several patients had to be excluded for various reasons: one patient withdrew from the study, two discontinued therapy early due to side effects and seven patients were excluded because of the presence of a stoma or pouch (since these fecal samples are not representative of the content of the whole intestinal tract). Additionally, five baseline samples were not collected, four samples failed sequencing, and two samples had to be excluded due to a low number of sequencing reads. The samples from 79 patients from which all data layers were complete (taxonomy, pathways and metabolites), were used for analyses (**Figure 1A**).

**Figure 1:**
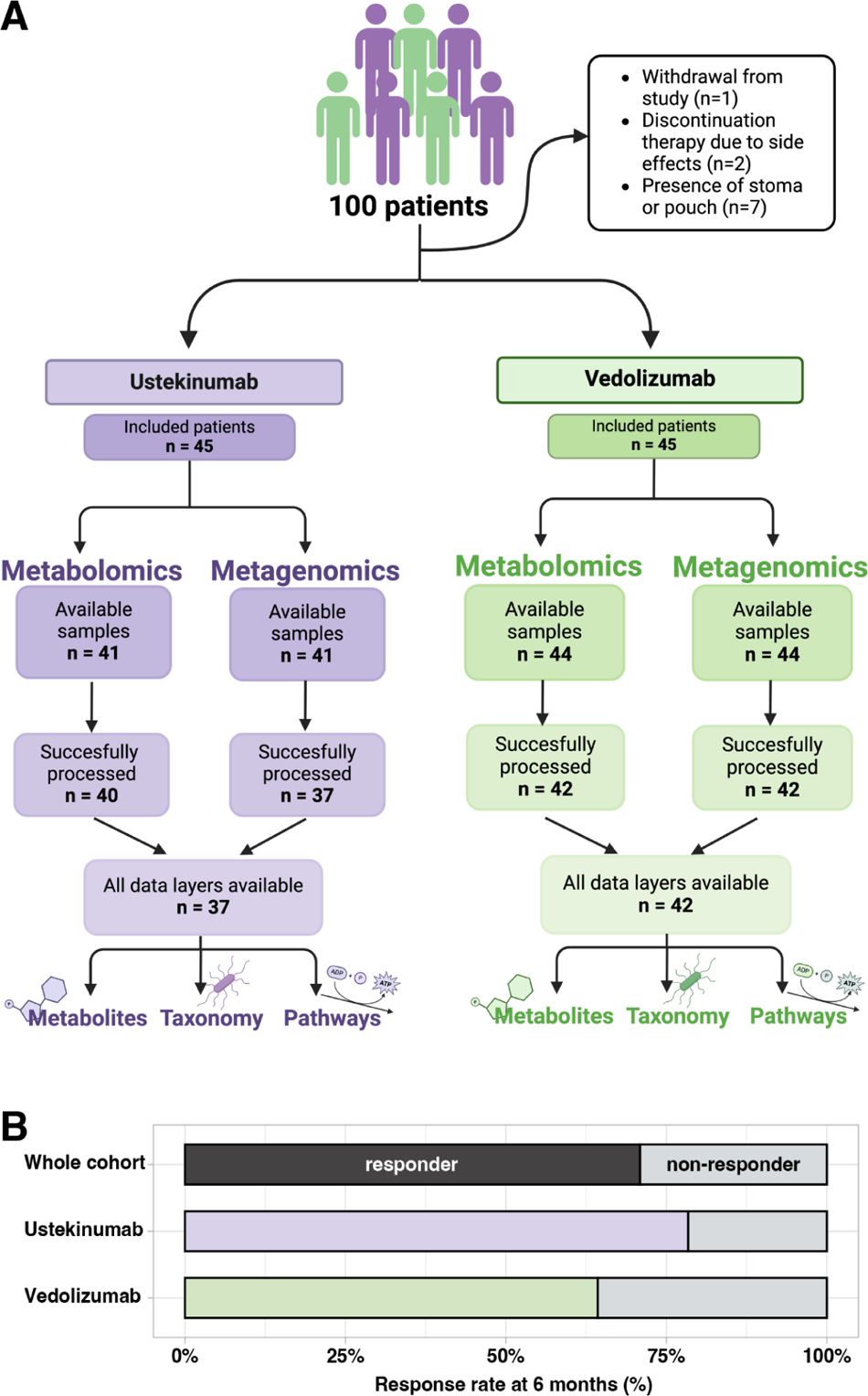
Cohort and sample overview A) Flowchart showing the available samples for the ustekinumab and vedolizumab group and the excluded samples. B) Responder and non-responders for the whole cohort and for ustekinumab and vedolizumab at 6 months after initiation of therapy.

### 1.2 Clinical characteristics

The study cohort consisted of 79 patients with IBD, initiating biologic treatment with vedolizumab (n=42) or ustekinumab (n=37). Fifty-nine percent of patients had CD, 29% UC and 11% of the patients IBD-U (**Table 1**). The sex distribution was slightly skewed towards females (54%). Among the participants, 23 patients were categorized as non-responders at 6 months, while 56 (71%) were responders (**Figure 1B**). Average age and BMI were comparable between the groups. Non-responders had higher clinical scores at baseline (SCCAI) than responders (p < 0.001). Additionally, levels of fecal calprotectin tended to be higher in non-responders (1856) at baseline compared to responders (1149), with the trending towards statistical significance (p = 0.07).

**Table 1:**
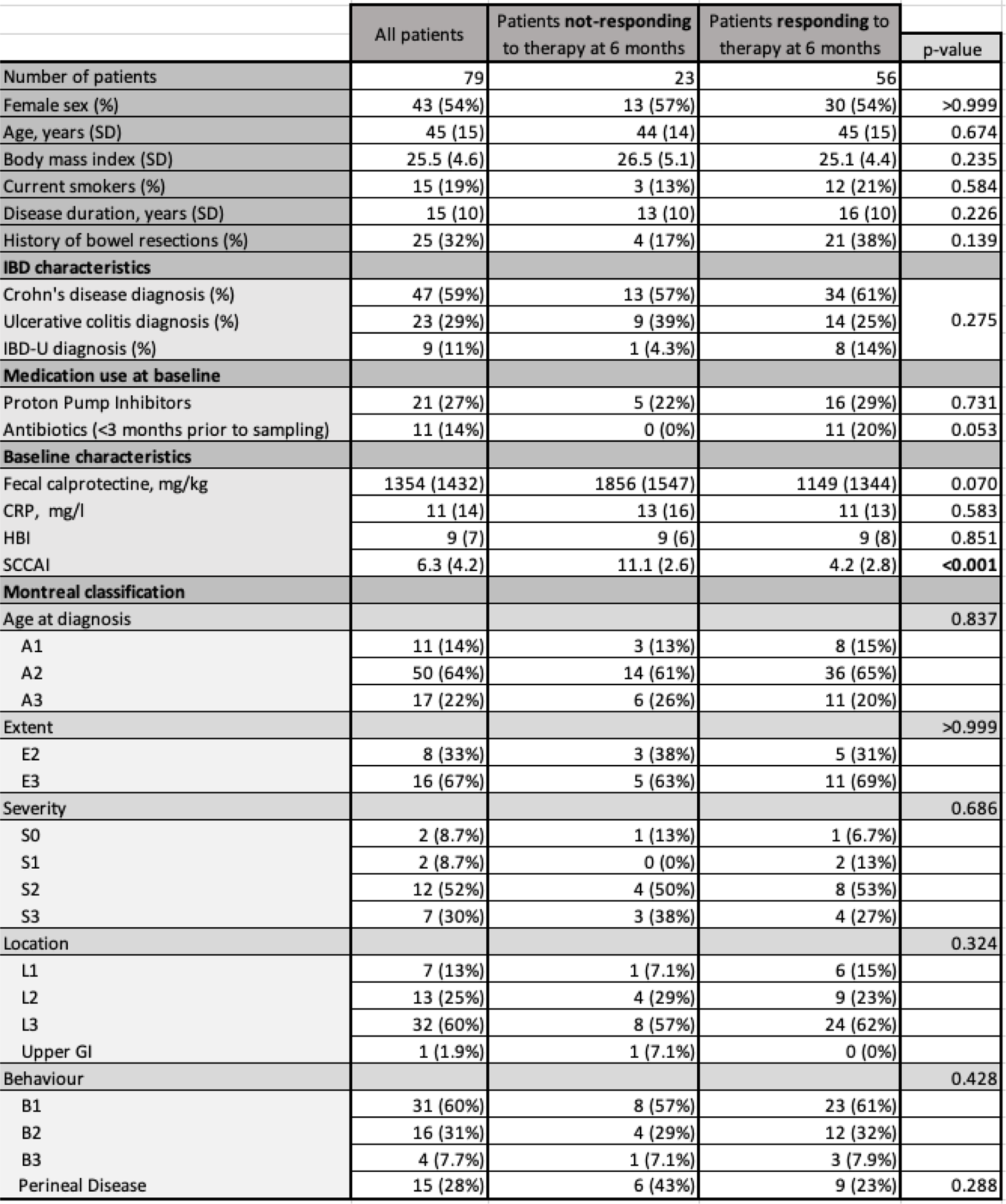
cohort description.

## 2. Baseline analyses of metagenomic and metabolomic data

### 2.1 Comparable baseline gut microbiome diversity and composition between responders and non-responders

We compared the gut microbiome between responders and non-responders at baseline, i.e. prior to the start of biologic treatment, to identify a possible microbiome signature that predicts response to biologics. We found no differences in baseline Shannon diversity between responders and non-responders (Mann-Whitney U test 0.56, Figure 2A), also when stratifying by biologic (ustekinumab p = 0.76, vedolizumab p = 0.64). No clustering in beta-diversity was observed between responders and non-responders in either the entire cohort or when stratified by biologic treatment, suggesting a comparable overall species composition (Figure 2B). When assessing the impact of responder status on the variance in microbial species composition through multivariable PERMANOVA, it did not explain a significant part of the variance (R2 = 0.008, p = 0.96). However, sequencing read depth (R2 = 0.025, p = <0.001), prior resections (R2 = 0.023, p = 0.002), and biologic (R2 = 0.023, p = 0.005) did contribute significantly to the composition variance. For pathways, responder status did not explain a significant portion of the variance as well, while antibiotic usage (R2 = 0.026, p = 0.047) and sequencing read depth (R2 = 0.032, p = 0.014) were found to have significant effects. This suggests that at baseline there is no distinction in the functional abilities of the gut microbiome between responders and non-responders, however the use of antibiotics does seem to affect the metabolic potential of the gut microbiome.

**Figure 2:**
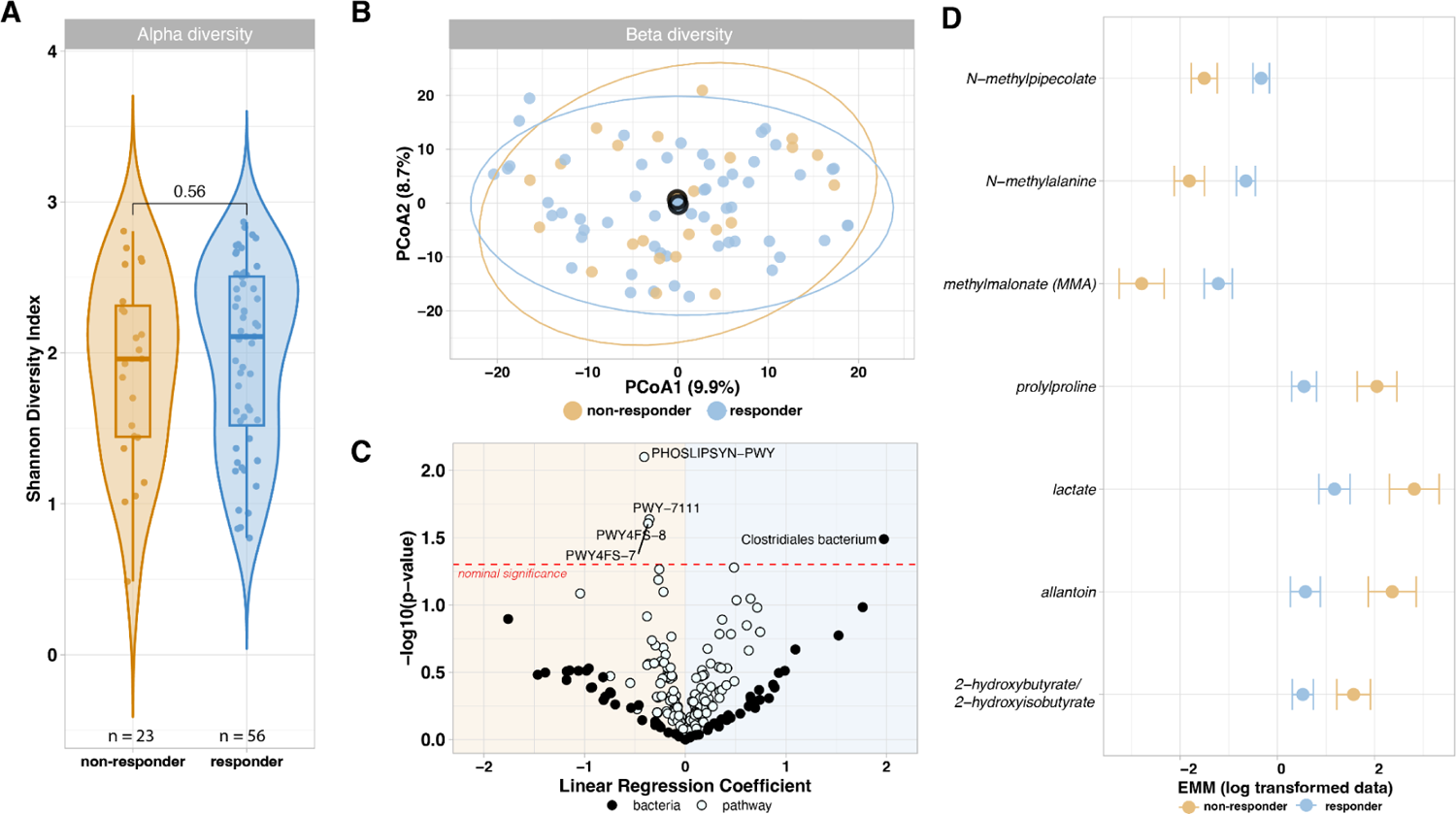
Baseline alpha diversity, beta diversity and differential abundant microbes, pathways and metabolites between responders and non-responders. Shown are the comparisons of 79 patients taking ustekinumab or vedolizumab, categorized by their response to therapy after 6 months. A) Alpha diversity between responders and non-responders displays no difference between these groups. (Mann-whitney U, p=0.56) B) Beta diversity between responders and non-responders using the Aitchinson distance. The overlapping centroids indicate no difference at the species level between responders and non-responders. C) Nominally significant p<0.05 relative abundant pathways and microbes between responders and non-responders. *Clostridales bacterium* and four pathways are associated with response, but these results do not pass the FDR <0.1 threshold. D) Seven metabolites showing significant differences (FDR <0.1) between responders and non-responders, suggesting that the only differences between responders and non-responders at baseline appear within the abundance of specific metabolites.

### 2.2 No differential abundant species and pathways between responders and non-responders

Next, we explored whether the microbiome composition at species and pathway level was correlated to response. We modeled CLR-transformed species and pathway abundances using linear regression, accounting for covariates as described in the methods. Our analysis did not identify any species with differential abundance between responders and non-responders at baseline that surpassed the FDR threshold of 0.1. However, when considering nominal significance, we observed an increase in the species *Clostridiales bacterium* in responders (p = 0.032, estimate = 1.973, Figure 2C). Stratifying the analysis by biologic treatment did not show any species reaching FDR significance. Our analysis of functional pathways revealed no FDR significant pathways with differential abundance between responders and non-responders at baseline, however when considering nominal significance (p<0.05) four pathways were found to be increased in non-responders (Figure 2C). These pathways are involved in phospholipid biosynthesis, pyruvate fermentation, and phosphatidylglycerol biosynthesis.

### 2.3 Baseline differences in metabolites between responders and non-responders

Next, we analyzed the metabolomics data with the hypothesis that responders might display a distinct metabolite profile compared to non-responders prior to the initiation of biologic treatment. We used the data of 816 metabolites present in at least 70% of the samples, applying linear regression to the natural-log transformed data. We identified seven metabolites with significant differential abundance at FDR level (Figure 2D), belonging to different metabolite classes. Upon stratification by biologic treatment, we did not observe any differentially abundant metabolites at baseline that reached FDR significance at <0.1. When only looking at bile acids, and grouping all available PBAs (n = 15) and SBAs (n = 43), we found that the logratio of PBA/SBA showed a significant difference between responders and non-responders (p = 0.035, estimate −1.233).

### 2.4 Network analysis shows interactions between microbial and metabolite clusters but limited associations with response

While our differential abundance analysis showed limited to no differences in gut microbiome features between responders and non-responders, it is important to take into account that this method focuses on isolated features. However, the microbiome is a complex ecosystem of microbes interacting with each other. Although the effects of each feature might be small or seem insignificant on its own, investigating them as part of a community considering the interplay between microbes (and metabolites), might reveal interesting patterns. Therefore, to explore the association between microbes and metabolites and test correlations with treatment response, we used *MiMeNet*. We identified seven microbial clusters and ten metabolite clusters. Figure 3 displays each cluster, indicating the number of features per cluster. While some example microbes and metabolites are shown per cluster, it is important to note that clustering was based on abundance rather than biological function or relevance, so the labels may not represent each cluster accurately. We observed that the cluster containing *Faecalibacterium prausnitzii* and the cluster containing *Alistipes finegoldii* showed opposite associations with metabolite clusters compared to the cluster containing *Escherichia coli.* Both *Faecalibacterium prausnitzii* and *Alistipes finegoldii* are generally associated with health. Next we aimed to associate these distinct microbiome-metabolite clusters with response. Three clusters were associated with responders; specifically, microbiome cluster 3 (Mann Whitney U, p=0.017) containing *Clostridiales bacterium*, and metabolite cluster 9 (Mann Whitney U, p=0.0499) containing urate, were positively associated with response to biologic therapy. On the other hand, we found metabolite cluster 5 (Mann Whitney U, p=0.01) containing many ethanolamides, to be positively associated with non-response to biologic therapy.

**Figure 3:**
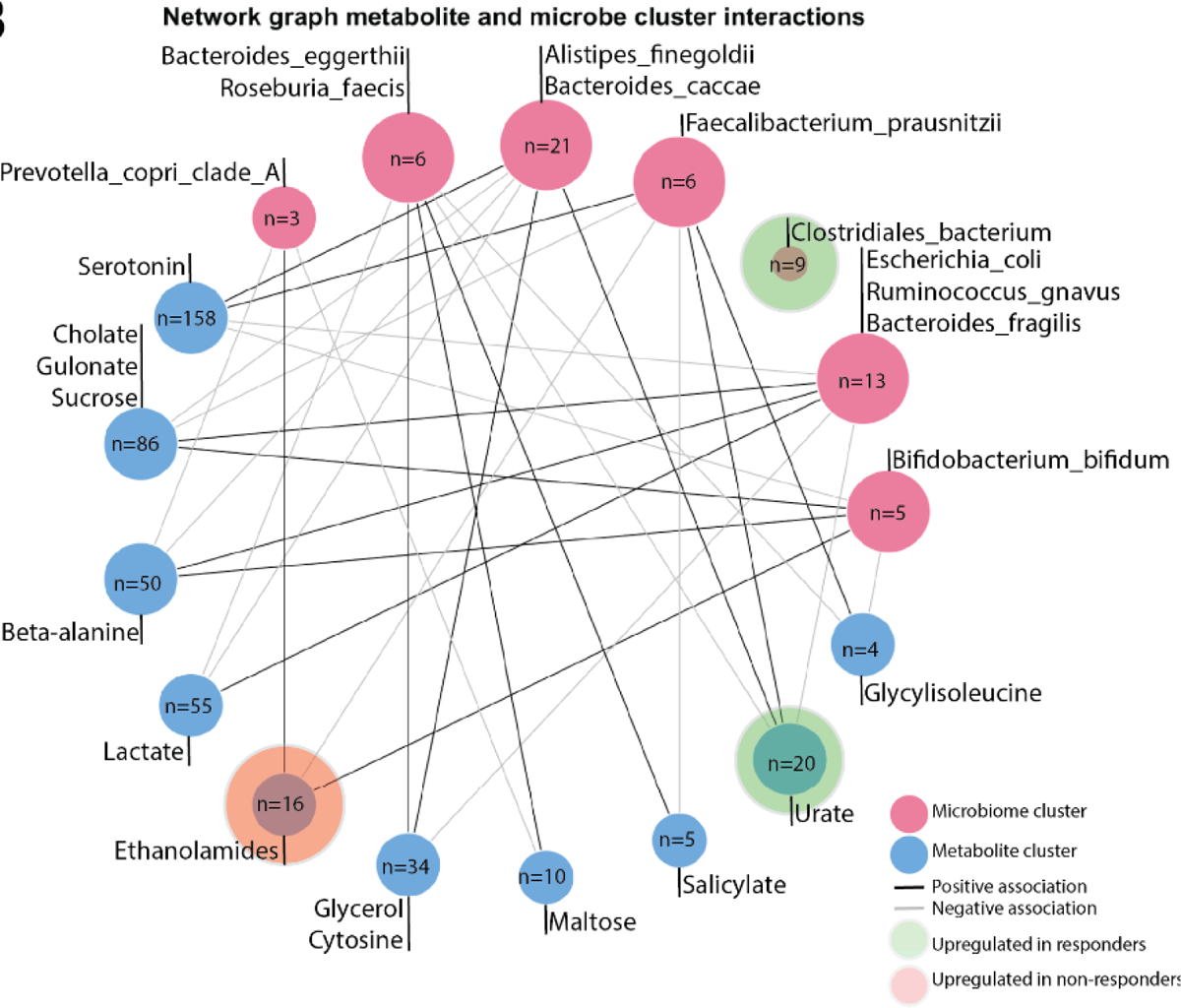
Microbiome-Metabolite interactions Network plot showing the interaction between microbial clusters and metabolite clusters in the whole cohort for 79 patients treated with Vedolizumab and Ustekinumab, created using MiMeNet. Clusters are based on co-occurrence, not biological relation; a full overview of features per cluster is available in the Supplemental Table S1 and S2. For each cluster one or more metabolites and bacteria are highlighted based on potential relevance. This representative does not have statistical or biological ascendancy over any other species or metabolite in the cluster. 2 metabolite clusters were significantly associated with response (Mann Whitney U, p=0.0499, p=0.01). And one bacterial cluster was significantly associated with response (Mann Whitney U, p=0.017).

## 3. Prediction of response to biological therapy

### 3.1 Limited predictive power for treatment response of metagenomic and metabolomic data

Following the more complex modeling approach, we implemented the CoDaCoRe package to determine the predictive power of ratios between metabolites, microbes or pathways in our dataset. Because of the limited sample size, we observed large variation based on which samples were randomly assigned to the test and training sets, therefore we chose to run 100 permutations of this assignment and averaged the AUCs across these permutations. In the combined cohort we observe an average test AUC of 0.59±0.09 for MGS species and 0.58±0.07 for the metabolites. When stratifying the prediction for each biological therapy, we observed test AUCs of 0.78±0.15 for each microbiome and metabolite predictors independently in the ustekinumab cohort, and 0.65±0.14 and 0.66±0.12 for microbiome and metabolites respectively in the vedolizumab cohort, indicating a difference in predictive power in microbial or metabolite features or response mechanism between vedolizumab and ustekinumab. The predictors are shown in Supplemental Figure S1. Because the individual cohorts have limited power for prediction we focused on the combined cohort. The features used in the prediction are shown in Figure 4A. An overview of each independent log ratio is shown in Figures 4B-D with the combined ratio shown in Figure 4E.

**Figure 4:**
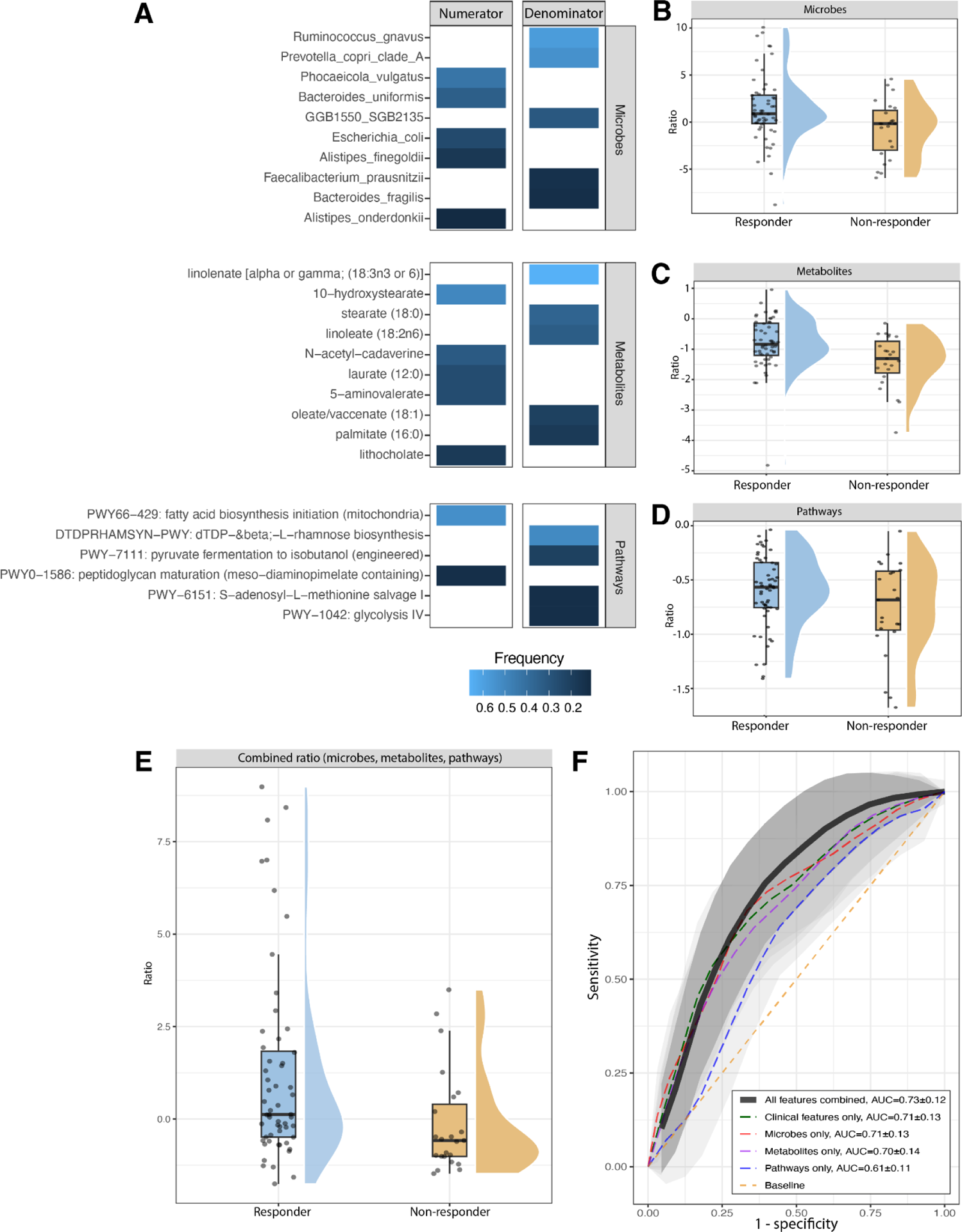
Features used in the prediction models, visualized feature ratios and prediction AUC overview A) Performing permutation analysis for the CoDaCoRe feature selection generated features for each of the categories, shown is the features with a frequency of 10% or higher. Stronger predictors are observed with a higher frequency. B, C, D) Visualized log ratios using the features in panel A from the abundances of the whole cohort data, shown are microbial, pathway and metabolites ratios. Densities of the responders and non-responders show limited separation. E) Combined plot of the ratios visualized in B,C and D. Responders are higher on average, although the largest density area still overlaps. F) ROC-AUC plot showing the AUC for the Generalized Linear Models based on clinical features, and microbial, pathway and metabolite ratios independently, and combined into one model. AUCs were determined using 100 permutations of 75% test and train split. Clinical features showed the best performance for each of the individual predictions, and combining multi-omic predictions only improved the prediction marginally.

To test the performance of the models created with the CoDaCoRe package we compared a model based purely on clinical features, and each ratio alone, and then the clinical features combined with the ratios. We observed similar predictive power from the clinical features alone, the microbiome ratios alone and the metabolite ratios alone, AUCs of 0.71±0.13, 0.71±0.13 and 0.70±0.14. Only pathways showed less predictive power at 0.61±0.11. Combining all ratios and clinical features only marginally improved the prediction to 0.73±0.12. Comparing the model fit between the model containing only clinical features and the model containing all three ratios in addition to the clinical features showed a marginally significant improvement (Likelihood ratio test, p=0.04986). All AUCs are shown in Figure 4F.

### 3.2 External cohort validation shows no predictive power of microbial features

To validate our findings, we included an external cohort from the University Medical Center Utrecht. Baseline fecal samples were collected and response status was determined by biologic continuation at six months of treatment. The Utrecht cohort comprised 47 participants: 10 started with adalimumab, 17 with infliximab, and 19 with vedolizumab. First, we selected the predictive microbe features for vedolizumab from our previous analysis and used them in the Utrecht vedolizumab patients (Figure 5A), when comparing this ratio plot with the plot shown in Figure 4B, the direction of the effect is reversed. Secondly, we compared the microbe features from the whole cohort with the anti-TNF Utrecht group, attempting to identify a generalized microbial signal indicating response to medication. This ratio is shown in Figure 5B, here the non-responders group cluster together around the lower end of the ratio, but the responders are seen across the full range. Testing these ratios in GLMs, we observed an AUC of 0.8 based on only clinical features in the vedolizumab cohort; additional inclusion of the microbiome ratio slightly worsened performance (Figure 5A). Testing a GLM in the anti-TNF cohort had worse performance compared to the vedolizumab cohort with an AUC of 0.655, which was also lowered slightly by the inclusion of the microbiome ratio to 0.639, although these values are very close together (Figure 5B), comparing these model fits also showed no significant improvement upon adding the microbiome ratio to either model (likelihood test, p>0.05). Overall, we observe limited to no predictive power of our models in the Utrecht cohort based on the microbial features identified in our cohort.

**Figure 5:**
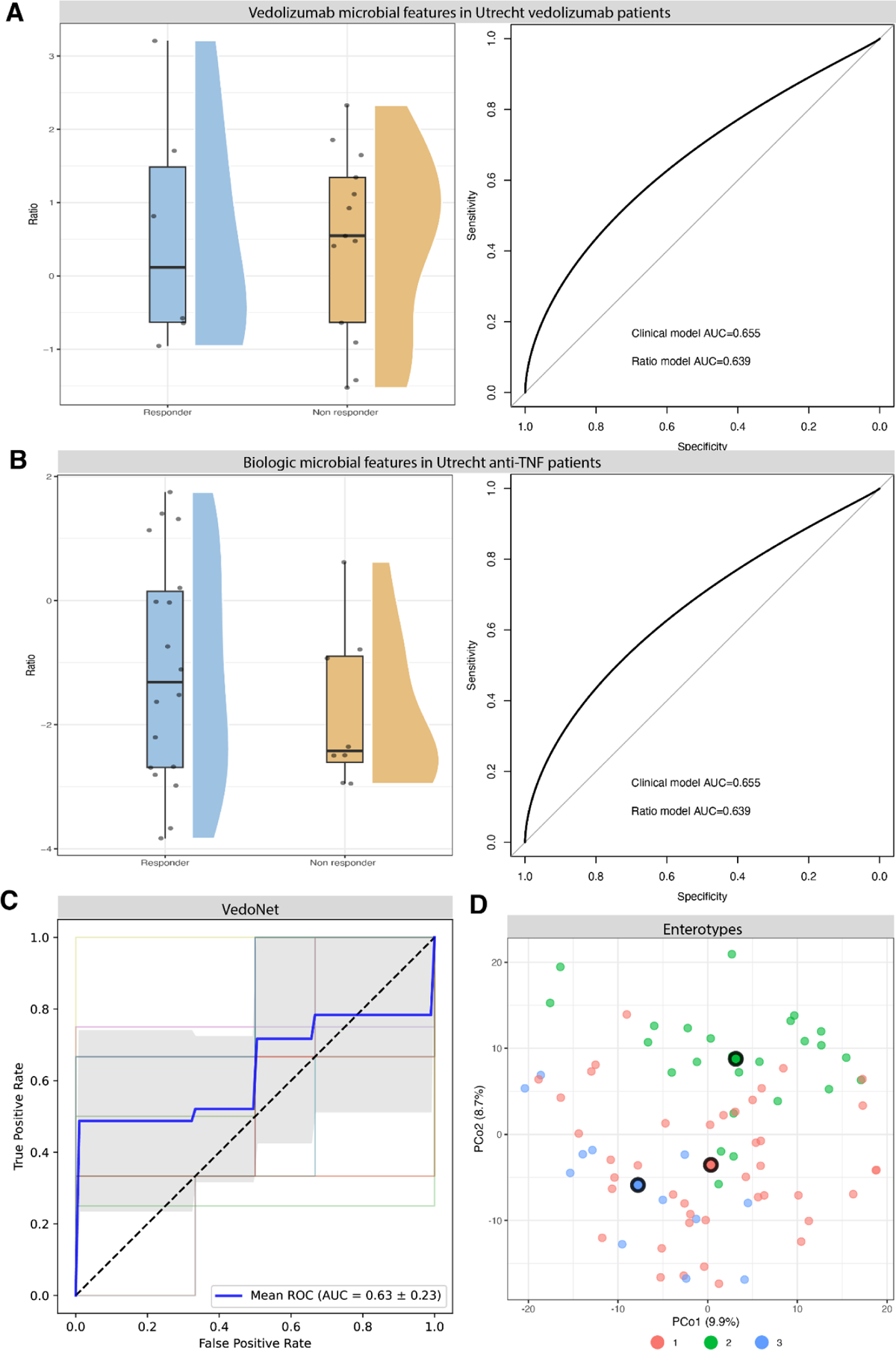
Utrecht validation and other prediction replication A) 6-month vedolizumab features plotted as a log ratio based on Utrecht cohort values and ROC-AUC plot from GLM models based on clinical features alone and clinical features plus 6-month vedolizumab microbiome features log ratio. Models were trained on the vedolizumab cohort and tested on the Vedolizumab patients in the Utrecht cohort. B) 6 month combined cohort features plotted as a log ratio based on Utrecht cohort values and ROC-AUC plot form GLM models based on clinical features and clinical features alone and clinical features plus 6 month combined cohort microbiome features log ratio. Models were trained on the combined cohort and tested on the anti-TNF patients in the Utrecht cohort. C) ROC-AUC curve based on Vedonet features in our vedolizumab cohort, determined using 4 fold 5 repeat k-fold cross validation in a random forest model. D) PCoA plot for enterotype clustering in our combined cohort.

## 4. Replication of previous published predictive microbiome features

### 4.1 VedoNet model features do not predict response in our dataset

In a previous study in 85 patients treated with vedolizumab, the authors created a prediction model, VedoNet, consisting of a selected set of 54 microbiome and clinical variables, which demonstrated strong discriminatory ability in predicting clinical remission (AUC = 0.872)^10^. We were able to match 49 out of the 54 features in our data and had complete information for each of these features for 22 Vedolizumab patients. Using these features of VedoNet, we aimed to predict response to vedolizumab using a random forest model. Our replication analysis resulted in an AUC of 0.63±0.23 (Figure 5C). Our prediction has no predictive power when compared to the reported AUC of 0.872, indicating that these features do not have the same predictive capability in our dataset.

### 4.2 Gut enterotypes do not associate with response

A previous study showed the predictive power of the Bact2 enterotype in combination with other stool factors^17^. Our attempt to repeat analysis using DMM revealed that the optimal number of dirichlet components based on Laplace or BIC approximation was three within our dataset, resulting in the identification of three distinct communities, i.e. enterotypes (Figure 5D). One community is defined by a high prevalence of *Prevotella*, the other two by a high prevalence of *Bacteroides*, with one of the *Bacteroides* communities also featuring a high prevalence of *Faecalibacterium*. Incorporating additional samples from the DMP to the abundance matrix resulted in the identification of two clusters (one with a high abundance of *Prevotella copri*), aligning with previous findings reported by Gacesa et al^18^. Next, we examined potential associations between enterotypes and treatment response outcomes. The three identified enterotypes were included in a Chi-square test, which did not display a significant association between enterotypes and response (X^2^ (2, N = 79) = 1.59, p = 0.45).

In a recent study, the predictive potential of enterotypes, specifically *Bacteroides2* (Bact2), in vedolizumab response was highlighted^17^. Bact2 is characterized by a depletion of butyrate-producing bacteria, reduced microbial load, and is associated with gastrointestinal inflammation. We identified a *Bacteroides* community in our cohort that shares similarities with the Bact2 enterotype, showing low abundances of butyrate-producing bacteria and reduced richness compared to our other communities. We investigated the relationship between this enterotype and response to vedolizumab and ustekinumab in our cohort using logistic regression. Our analysis did not identify any significant association between response and our Bact2 (coefficient −0.2, p = 0.70). Additionally, we tested response and other baseline variables, and found that previous anti-TNF use is significantly correlated to response to ustekinumab or vedolizumab (coefficient = 1.31, p = 0.027, n=18 anti-TNF naive).

## 5. Response definitions based on short- and long-term response show different results

Finally, we repeated all analyses to identify features associated with short term response – based on HBI/SCCAI scores at week 14 and 16 and long-term response/durable response. Durable or prolonged response, defined as continuation of therapy after ∼2 years, occurred in 48 patients (55%), while 40 patients discontinued therapy, indicating that 17 patients experienced a loss of response between six months and ∼2 years after initiation of treatment. In two patients the response after ∼2 years could not be confirmed because of relocation to another part of the country or their passing. Similarly, defining response using clinical scores resulted in another distribution, with 35 responders (42%) and 48 non-responders identified.

Diversity and dissimilarity analyses were conducted for the additional definitions of response, but no baseline differences between responders and non-responders were observed for either definition. Subsequently, we performed differential abundance analysis for microbes, pathways and metabolites based on each definition of response, considering the total cohort and stratifying by biologic treatment. Interestingly, the majority of features showing differential abundance in one response-definition-cohort combination were specific to that combination. There was limited overlap in features between the same definition across the three various cohorts, and similarly there was limited overlap between definitions within the same cohort (Figure 6).

**Figure 6:**
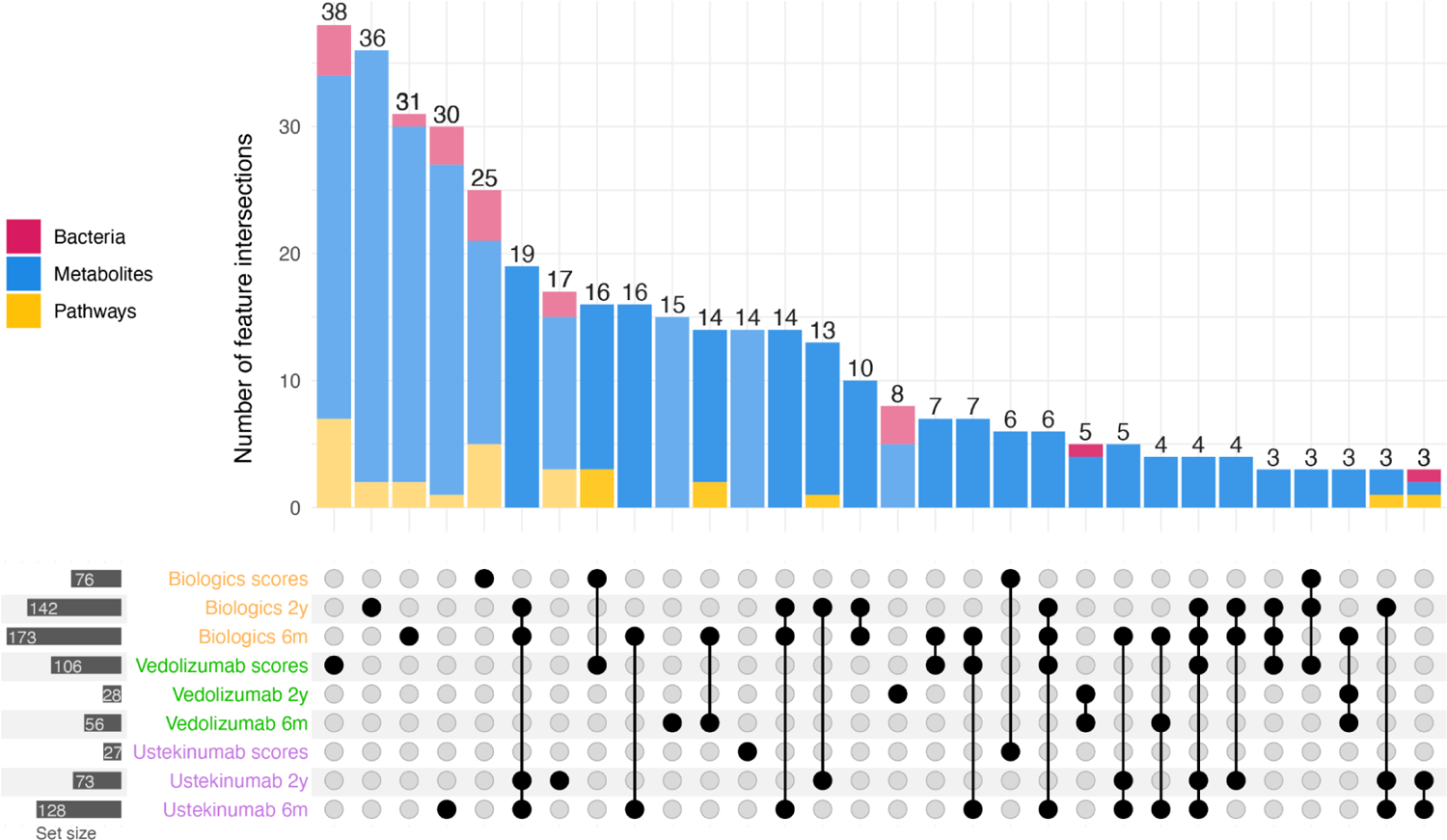
Overlapping features based on three response definitions UpSet plot showing the number of features overlapping between nine different sets: the entire cohort and the cohort stratified by biologic, and the three definitions of response. The categories of the overlapping features are indicated in pink (bacteria), blue (metabolites) and yellow (pathways).

## Discussion

In this prospective study our aim was to identify factors of the gut microbiome predictive of response to biologics (vedolizumab and ustekinumab) in patients with IBD. Several studies have linked baseline microbiome composition to therapy outcomes, suggesting that the gut microbiome plays a critical role as a potential modulator and predictor of treatment response^7,10,11,17^. We took a comprehensive approach and considered clinical characteristics, gut microbiome species, pathways and fecal metabolites. Our analyses, focusing on the alpha and beta diversity of the gut microbiome at baseline, both at species level and pathways, did not reveal significant differences that could help to identify future responders or non-responders. Seven metabolites were found to differ at baseline between responders and non-responders. In line with the minimal differences in gut microbiome and metabolite profiles at baseline in relation to treatment response, the AUCs of our prediction analysis indicated that a combination of these features (AUC=0.73±0.12) might only marginally aid in predicting response to biologic treatments. While our cohort is one of the largest reported to date, the added predictive power of microbiome features was limited, implying a lack of biological effects from the microbiome on the outcomes of ustekinumab and vedolizumab therapy.

Previous studies have explored the relationship between the gut microbiome and its metabolites, and outcomes of biologics treatments in IBD, focusing particularly on its predictive capabilities. In line with prior studies, we found some predictive power associated with higher abundance of specific secondary bile acids, like lithocholic acid, in our models^15,19^. However, we did not find a difference in abundance of this SBA between responders and non-responders. Interestingly, when grouping the available PBAs and SBAs in our dataset, the logratio PBA/SBA was significantly different between responders and non-responders. We could not further investigate these signals in our replication cohort as there were no fecal metabolites available. Additionally, a study reported that CD patients responding to vedolizumab showed significantly higher alpha-diversity at baseline, although this difference did not reach significance in UC patients^10^. We did not identify baseline differences in alpha-diversity between responders and non-reponders. Furthermore, they also found that PCoAs failed to differentiate between remitters and non-remitters, which aligns with the findings from our study. While they identified species (*Roseburia inulinivorans* and *Burkholderiales*) to be more abundant at baseline in CD remitters compared to non-remitters, these species were not identified as differentially abundant in our analyses. Also, their predictive model, termed VedoNet (AUC=0.872), did not show the same predictive power in our vedolizumab cohort (AUC=0.61). However, it is worth noting that there are several disparities between the studies, potentially explaining the lack of overlap in results: 1) differences in geographic origin of the patients, including differences in lifestyle 2) different definitions of endpoints (remission at 14 weeks based on disease severity score versus our 6 month response definition), 3) fecal sample collection and processing differences and 4) differences in health care systems, which may lead to different patient populations that get treated with vedolizumab. In another study, it was demonstrated that a model based on clinical data, stool characteristics and the Bact2 enterotype showed predictive power for treatment outcomes (73.9% accuracy for biological therapies)^17^. Interestingly, their model including only clinical data, showed a comparable ROC-AUC value of 73.5% Our study findings show a similar pattern. Our model including microbiome features and clinical features only marginally improved the clinical model, suggesting that the clinical model alone hold considerable predictive value and inclusion of microbiome factors only slightly improves the predictive capability.

The absence of baseline differences in microbial diversity between responders and non-responders before the start of treatment may be attributed to the diverse disease courses and prior management strategies among the patients. Patients starting with biologic treatment often have a history of exposure to an intense range of therapies, for instance, 80% of our patients had prior exposure to TNFalpha antagonists. This exposure could induce substantial alterations in both the gut microbiome and metabolite profiles prior to sampling. Consequently, this may result in challenges to distinguish significant differences between responders and non-responders, i.e. taxa, pathways or metabolites in a cohort of this size. Furthermore, given the well-established fact of high heterogeneity in gut microbiome composition between individuals^20^, the search for broad indicators or ‘biomarkers’ of the gut microbiome may give results that are small and too nuanced. The individual’s optimal microbiome composition for response most likely varies, underscoring the need for personalized medicine.

Our analyses underscore the significant impact of the chosen definition of response and the timing of defining response on study outcomes. Definitions relying on disease severity scores (HBI and SCCAI) possess a subjective nature. They might capture symptoms resembling and belonging to irritable bowel syndrome rather than accurately reflecting active disease of IBD. This study had the opportunity to use extensive information captured by the medical records of the patients. We believe that the optimal representation of the clinical context involves a broad approach, including a combination of clinical disease activity scores, routine laboratory diagnostic values, fecal calprotectin and most importantly, the global assessment of the treating gastroenterologist based on these factors. For generalizability, our study could have benefitted from incorporating endoscopic response scores. However, it is important to acknowledge that our study was not a randomized trial, and patients received standard care. The standard care practice does not involve evaluation of endoscopic response.

To conclude, our study showed that within our prospective cohort of IBD patients undergoing treatment with ustekinumab or vedolizumab, no significant differences in the gut microbiome at baseline between response and non-responders were observed, and incorporating microbial or metabolite features did not improve predictive power, leading us to infer that the gut microbiome at this stage of IBD may have very little, or no impact on the outcome of treatment with either vedolizumab or ustekinumab. The lack of replication of other prediction methods suggests that predictive models, whether successful or not, appear limited to the initial study cohorts of each prediction. Additionally, the sample size and sparsity of microbiome datasets in many studies increases the risk of overfitting and false positives, emphasizing the importance of external validation. Based on our findings and efforts to identify and replicate predictors for treatment response, it appears that predictors of therapeutic success are not found in the fecal microbiome.

## Data Availability

All data produced in the present study are available upon reasonable request to the authors

## Acknowledgments

The authors would like to thank all study participants for fecal sample collection. Also, we thank I. Tamargo Rubio for help designing the figures. Figure 1 was created using BioRender.com

## Contributors

FMP and IJH contributed equally to this work. MAYK and VC also contributed equally. Conceptualization: MAYK, VC, RK. Data collection: JPDS, WTCUV, AM, JBA, WGNM, BJMW. Analysis and interpretation of the data: FMP, IJH. Writing - original draft: FMP, IJH, MAYK, VC. Writing - reviewing: all authors. Supervision: HHF, BO, RG, RK

## Funding

The study received financial support through Investigator Initiated Study Grants from Johnson & Jonson and Takeda Pharmaceuticals.

## Conflicts of interest

EAMF is supported by a ZonMW Clinical Fellowship grant (project number 90719075) and has received an unrestricted research grant from Takeda. RG received funding by Janssen Pharmaceuticals (for unrelated research projects) and received consulting funding from Esox Biologics (for unrelated research projects). RKW has received unrestricted Research Grants from Takeda, Johnson & Johnson, Ferring and Tramedico and speakers fees from Abbvie, MSD and Boston Scientific and has acted as a consultant for Takeda Pharmaceuticals.

---

**Supplementary Figure S1.**
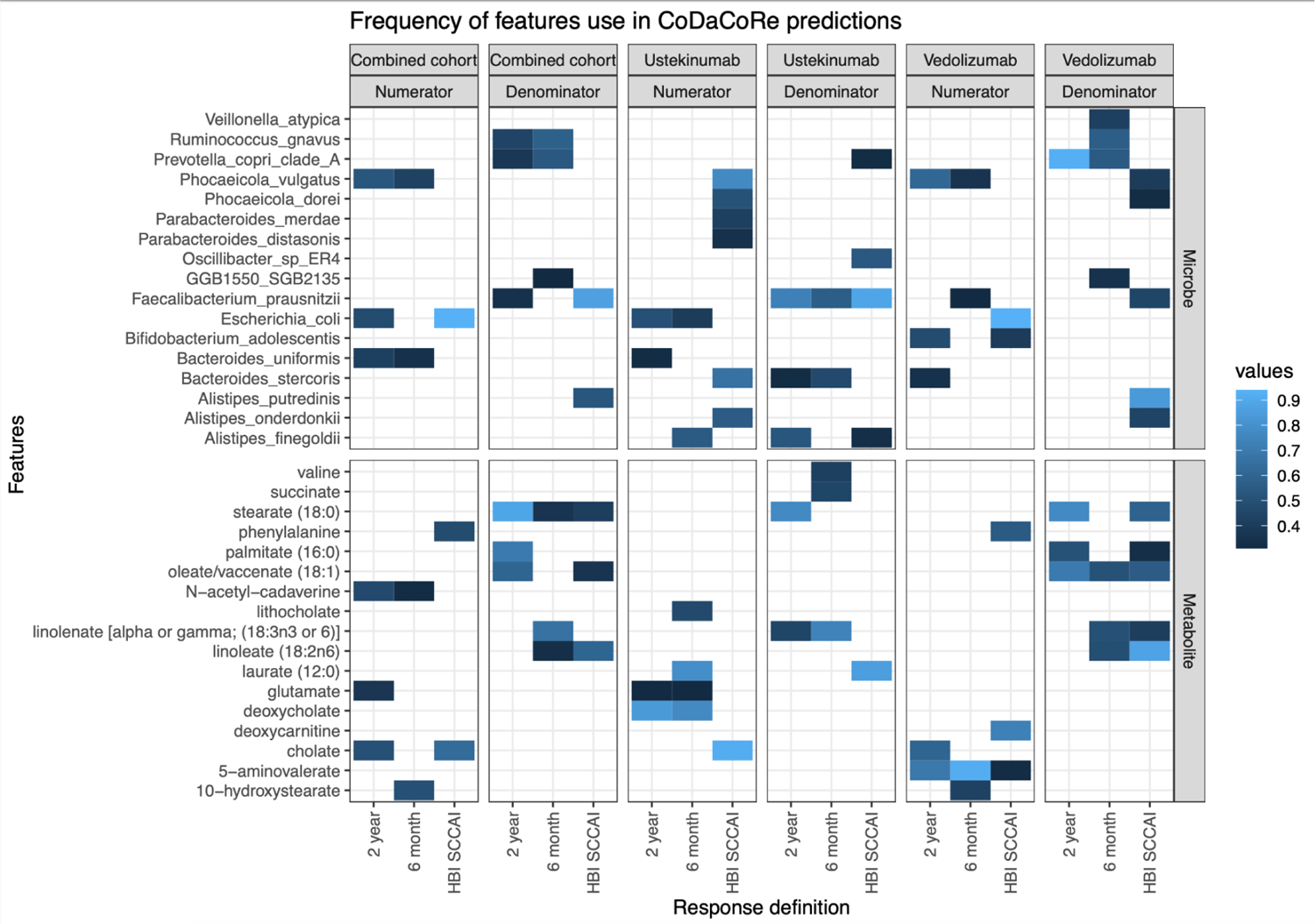
Frequency of features used in the CoDaCoRe predictions over all medication and response definitions Shown are the predictors from the CoDaCoRe permutation analysis. Only features selected more than 10% of the time are shown. For each cohort the Numerator and Denominator are shown and for each response definition. Features highlighted in the numerator are more abundant in responders and features in the denominator are more abundant in non-responders.

Supplementary table S1

MiMenet metabolite features

See attached excel file

Supplementary table S2

MiMenet microbiome features

See attached excel file

## Notes

### Author Declarations

IRB University Medical Center Groningen Netherlands Consent from all participants was obtained through GEID (NL58808.042.16) andr Parelsnoer IRB (NL24572.018.08).

## References

1. Zhao M, Gönczi L, Lakatos PL, Burisch J. The Burden of Inflammatory Bowel Disease in Europe in 2020. J Crohns Colitis. 2021;15(9):1573–1587. doi:10.1093/ecco-jcc/jjab029

2. Nishida A, Inoue R, Inatomi O, Bamba S, Naito Y, Andoh A. Gut microbiota in the pathogenesis of inflammatory bowel disease. Clin J Gastroenterol. 2018;11(1):1–10. doi:10.1007/s12328-017-0813-5

3. Honap S, Meade S, Ibraheim H, Irving PM, Jones MP, Samaan MA. Effectiveness and Safety of Ustekinumab in Inflammatory Bowel Disease: A Systematic Review and Meta-Analysis. Dig Dis Sci. 2022;67(3):1018–1035. doi:10.1007/s10620-021-06932-4

4. Schreiber S, Dignass A, Peyrin-Biroulet L, et al. Systematic review with meta-analysis: real-world effectiveness and safety of vedolizumab in patients with inflammatory bowel disease. J Gastroenterol. 2018;53(9):1048–1064. doi:10.1007/s00535-018-1480-0

5. Wong DJ, Roth EM, Feuerstein JD, Poylin VY. Surgery in the age of biologics. Gastroenterol Rep. 2019;7(2):77–90. doi:10.1093/gastro/goz004

6. Gisbert JP, Chaparro M. Predictors of Primary Response to Biologic Treatment [Anti-TNF, Vedolizumab, and Ustekinumab] in Patients With Inflammatory Bowel Disease: From Basic Science to Clinical Practice. J Crohns Colitis. 2020;14(5):694–709. doi:10.1093/ecco-jcc/jjz195

7. Kolho KL, Korpela K, Jaakkola T, et al. Fecal Microbiota in Pediatric Inflammatory Bowel Disease and Its Relation to Inflammation. Off J Am Coll Gastroenterol ACG. 2015;110(6):921. doi:10.1038/ajg.2015.149

8. Zhou Y, Xu ZZ, He Y, et al. Gut Microbiota Offers Universal Biomarkers across Ethnicity in Inflammatory Bowel Disease Diagnosis and Infliximab Response Prediction. mSystems. 2018;3(1):e00188–17. doi:10.1128/mSystems.00188-17

9. Shaw KA, Bertha M, Hofmekler T, et al. Dysbiosis, inflammation, and response to treatment: a longitudinal study of pediatric subjects with newly diagnosed inflammatory bowel disease. Genome Med. 2016;8(1):75. doi:10.1186/s13073-016-0331-y

10. Ananthakrishnan AN, Luo C, Yajnik V, et al. Gut Microbiome Function Predicts Response to Anti-integrin Biologic Therapy in Inflammatory Bowel Diseases. Cell Host Microbe. 2017;21(5):603–610.e3. doi:10.1016/j.chom.2017.04.010

11. Doherty MK, Ding T, Koumpouras C, et al. Fecal Microbiota Signatures Are Associated with Response to Ustekinumab Therapy among Crohn’s Disease Patients. mBio. 2018;9(2):e02120–17. doi:10.1128/mBio.02120-17

12. Lavelle A, Sokol H. Gut microbiota-derived metabolites as key actors in inflammatory bowel disease. Nat Rev Gastroenterol Hepatol. 2020;17(4):223–237. doi:10.1038/s41575-019-0258-z

13. Vich Vila A, Hu S, Andreu-Sánchez S, et al. Faecal metabolome and its determinants in inflammatory bowel disease. Gut. 2023;72(8):1472–1485. doi:10.1136/gutjnl-2022-328048

14. Lee JWJ, Plichta D, Hogstrom L, et al. Multi-omics reveal microbial determinants impacting responses to biologic therapies in Inflammatory Bowel Disease. Cell Host Microbe. 2021;29(8):1294–1304.e4. doi:10.1016/j.chom.2021.06.019

15. Ding NS, McDonald J a. K, Perdones-Montero A, et al. Metabonomics and the Gut Microbiome Associated With Primary Response to Anti-TNF Therapy in Crohn’s Disease. J Crohns Colitis. 2020;14(8):1090–1102. doi:10.1093/ecco-jcc/jjaa039

16. Christensen KR, Ainsworth MA, Skougaard M, et al. Identifying and understanding disease burden in patients with inflammatory bowel disease. BMJ Open Gastroenterol. 2022;9(1):e000994. doi:10.1136/bmjgast-2022-000994

17. Caenepeel C, Falony G, Machiels K, et al. Dysbiosis and Associated Stool Features Improve Prediction of Response to Biological Therapy in Inflammatory Bowel Disease. Gastroenterology. 2024;166(3):483–495. doi:10.1053/j.gastro.2023.11.304

18. Gacesa R, Kurilshikov A, Vich Vila A, et al. Environmental factors shaping the gut microbiome in a Dutch population. Nature. 2022;604(7907):732–739. doi:10.1038/s41586-022-04567-7

19. Lloyd-Price J, Arze C, Ananthakrishnan AN, et al. Multi-omics of the gut microbial ecosystem in inflammatory bowel diseases. Nature. 2019;569(7758):655–662. doi:10.1038/s41586-019-1237-9

20. Ursell LK, Metcalf JL, Parfrey LW, Knight R. Defining the human microbiome. Nutr Rev. 2012;70(suppl_1):S38–S44. doi:10.1111/j.1753-4887.2012.00493.x

